# An Affordable Smartphone-based Fundus Imaging Device for Retinal Examination

**DOI:** 10.1101/2025.08.20.25333845

**Authors:** Solomon Gebru Abay, Melkamu Hunegnaw Asmare, Lucca Geurts

## Abstract

Fundus imaging is widely used for diagnosing and monitoring ocular diseases such as diabetic retinopathy, glaucoma, and age-related macular degeneration. However, the high cost and limited availability of conventional fundus cameras pose significant barriers, particularly in resource-constrained settings. This study introduces the Glaucoma Screening on Phone (GSoP), an affordable and portable smartphone-based fundus imaging adapter designed to address these challenges. The adapter was developed using accessible and cost-effective components. We recorded retinal videos from dilated pupil with a focus on the optic disc region, which provides critical information on the degeneration of the optic nerve. While clinical trials revealed artifacts such as glare and noise that reduced overall image quality, the GSoP demonstrated its ability to capture diagnostically relevant images of the optic disc region. A subjective qualitative comparison with the commercially available ophthalmoscope called oDocs-nun showed that although the GSoP’s field of view is smaller, it effectively highlights the optic nerve head, a critical area for glaucoma screening. Our approach is well-suited for mydriatic video-based screening due to its limited field of view. With a production cost of under C10, the GSoP offers a practical and accessible solution for primary healthcare and educational purposes. Future improvements, including glare reduction mechanisms, AI-driven automation, and modular design options, have the potential to enhance its diagnostic capabilities and broaden its impact.

## I. INTRODUCTION

Retinal diseases, such as glaucoma, age-related macular degeneration, diabetic retinopathy, and trachoma, are major contributors to global blindness and vision impairment [1], [2]. According to the World Health Organization, approximately 2.2 billion people worldwide live with some form of vision impairment, with at least 1 billion of these cases considered preventable [3]. Early detection of these conditions is vital, as it allows for timely intervention and management, significantly reducing the risk of irreversible vision loss. Detecting retinal abnormalities typically involves assessing the condition of the retinal wall through fundus images, which serve as a key component of the screening process to identify early signs of disease. However, access to traditional fundus cameras remains a significant barrier to widespread screening as these devices are often prohibitively expensive and typically bulky and non-portable, making them impractical for deployment in rural or resource-limited areas [4]. This lack of access to diagnostic equipment in underserved regions means that many individuals do not receive the essential care they need. Consequently, there is an urgent demand for affordable and portable retinal imaging solutions that can bridge this gap, enabling more widespread access to early detection and improved patient outcomes in communities that need it the most.

Recent advancements in technology have enabled smartphones to emerge as a viable alternative to many traditional, expensive medical devices [5], [6]. Smartphone-based fundus imaging systems capitalize on the computational power and high-resolution cameras of modern smartphones, offering a cost-effective and portable solution for retinal disease screening [7], [8]. These devices are increasingly being utilized for retinal screenings, particularly in developing countries, where approximately 80% of vision-impaired individuals live in remote areas with limited access to healthcare [9], [3]. By simplifying the screening process, smartphone-based systems eliminate the need for expensive equipment and reduce the requirement for extensive operator training. This makes them an ideal solution for primary care settings and resource-limited environments, where access to specialized medical devices is often restricted [10], [11]. Furthermore, their portability allows for mobile screening efforts, enabling healthcare workers to perform fundus examinations in rural, underserved regions, thus reaching populations that would otherwise remain undiagnosed. The integration of these systems into existing healthcare workflows holds the potential to significantly improve early detection rates and facilitate timely interventions, ultimately reducing the burden of preventable vision impairment.

Devices such as the Vista View from Volk [12], D-Eye [13], and Remidio Fundus On Phone [14] have demonstrated the feasibility of capturing high-quality retinal images, even in challenging settings such as low-resource healthcare settings, environments with limited lighting control, and situations where patient movement or lack of pupil dilation may affect imaging quality. These devices offer performance comparable to standard fundus ophthalmoscopy machines, making them valuable tools for retinal diagnostics. However, they remain largely inaccessible in developing countries due to their cost, which, while lower than standard devices, is still prohibitively expensive for widespread use in resource-constrained settings. This study addresses this challenge by introducing a significantly more affordable smartphone-based fundus imaging adapter. Designed for use in rural areas, this device is ideal for screening purposes, providing a practical and accessible solution to bridge the gap in retinal care.

This paper introduces the Glaucoma Screening on Phone (GSoP), an affordable and portable smartphone-based fundus imaging adapter designed for glaucoma screening. The GSoP is developed using cost-effective and accessible components, and it is suitable for video-based screening. Key contributions of this study include:

- Development of a low-cost optical adapter that utilizes a smartphone’s camera to capture images of the optic nerve head (ONH) region. The device works best with mydriatic video recordings to compensate for its small FOV, and its production cost is significantly lower than the existing smartphone-based devices. This makes it more ideal to deploy on resource-constrained devices.
- Analysis of sample videos captured using the GSoP to assess the diagnostic quality of the images for glaucoma screening.
- Qualitative comparison of the GSoP with the oDocsnun ophthalmoscope (by oDocs Eye Care Limited, New Zealand) with respect to FOV and quality in capturing retinal videos.

The remainder of this paper is organized as follows: Section II reviews related works. Section III elaborates the design. Section IV presents the assessment of output images from our devices under results and discussion. Finally, Section V concludes the paper.

## II. Related works

Smartphone-based fundus imaging has gained significant attention as a cost-effective and accessible solution for diagnosing retinal diseases. Numerous studies have explored the potential of these devices, emphasizing their affordability, portability, and diagnostic accuracy.

An affordable and portable mydriatic fundus camera was developed by [15] through the modification of a Panasonic Lumix-G2 consumer camera with a custom optical module. The device incorporated low-cost optical components to provide uniform fundus illumination and a 50° retinal FOV. They produced fundus images of comparable quality to traditional tabletop fundus cameras, effectively capturing retinal pathologies such as diabetic retinopathy and agerelated macular degeneration. With a production cost under $1,000, the study highlighted its potential as a solution for retinal screening in resource-constrained environments, advancing accessible eye care.

The use of a 20D ophthalmic lens, which costs from $100 to $300, with a smartphone to capture fundus images was evaluated by [16] on dilated eyes. They developed an application to automatically select high-quality images from video frames, addressing the challenge of ensuring diagnostic-quality images. Similarly, [17] also recorded videos using a 20D lens and manually selected frames for analysis. While both approaches operated with pupil dilation similar to traditional fundus cameras, they also explored the feasibility of capturing fundus images without dilation, identifying challenges and future research directions.

The study by [18] introduced the Remidio Vistaro, a smartphone-based fundus imaging device designed for mydriatic use, costing just a tenth of the tabletop cameras. The device features an annular illumination system that reduces Purkinje reflexes and an autocapture algorithm that automates image acquisition upon achieving the correct working distance. With a single-shot FOV of around 65°, the device achieved clinically useful image quality in 91.6% of cases, and automatic image capture succeeded in 80% of examinations within 10–15 seconds. These findings highlight the potential of widefield smartphone-based imaging as a portable and accessible solution for detecting retinal pathologies.

A low-cost hardware attachment named the D-EYE adapter was designed by [13] for smartphones to capture still and live retinal images. Magnetically attached to the smartphone, this device was specifically designed for iPhone 5 and 6. Using the principles of direct ophthalmoscopy, it achieved clinical resolution with a FOV of approximately 20° on dilated eyes. Its cross-polarization optical design effectively reduced corneal reflections. The final prototype, measuring 47 × 18 × 10 mm, was portable and convenient. Despite its compact design, the adapter alone cost around $400, limiting its affordability in some settings.

A study by [19] developed a portable, low-cost, nonmydriatic fundus camera using a Raspberry Pi computer and readily available components, offering a FOV of 5 to 6 disc diameter. While not smartphone-based, their device utilized an infrared-sensitive camera board, a dual infrared and white light LED, a 20-diopter condensing lens, and a compact 5inch LCD touchscreen. Designed to fit in a white coat pocket, their prototype measured 133 mm × 91 mm × 45 mm, and cost $185.20 to produce, including the lens. By using the indirect ophthalmoscopy principles, their prototype captured good-quality fundus images on undilated pupil.

A glare-free nonmydriatic retinal imaging camera was introduced by [20]. The device utilized a Lytro Illum consumer light field camera and a unique optical design to optimize the relationship between the eye pupil, system aperture stop, and micro-image separation. This setup enhanced stereopsis compared to traditional stereo fundus cameras. The study also developed a glare identification and selective image rendering technique to eliminate corneal backscatter, resulting in high-quality, glare-free retinal images. Validated on physical models and live human eyes, the device demonstrated the capability to produce nonmydriatic color and 3D retinal images, offering a FOV of approximately 32 degree.

The reviewed studies highlights the potential of smartphone-based fundus imaging in addressing the limitations of traditional fundus cameras. These devices offer cost-effective, portable, and user-friendly solutions for retinal diagnostics, making them valuable in resource-constrained settings.

Building on these advancements, our study introduces the GSoP, a highly affordable smartphone-based fundus imaging device designed for glaucoma screening. By addressing cost barriers and emphasizing simplicity, the GSoP aligns with the broader goal of democratizing access to retinal diagnostics, particularly in underserved regions. With a total production cost of under C10, the GSoP is significantly cheaper than other smartphone-based fundus imaging devices and is also easily maintainable and sustainable, making it an ideal choice for rural screening initiatives and educational applications.

## III. Materials and Methods

### A. Design and Development

The GSoP consists of 3D-printed plastic components, a black sponge, a reflective mirror, transparent paper, and a beam splitter. The plastic components, including the main case and upper cover, were designed using Solid Edge 2022 (University Edition), as shown in Fig. 1. The device was intentionally designed for simple assembly, requiring minimal tools and expertise. The calibration process involved aligning the optical axis of the light emitted by the smartphone’s flashlight with the camera’s optical path to achieve optimal focus and FOV. The 3D model designs were exported in stereolithography file format and manufactured using a Prusa I3 MK3S 3D printer. The 3D-printed housing ensures accurate alignment and secure attachment of the optical components according to the specified measurements.

**Fig. 1.**
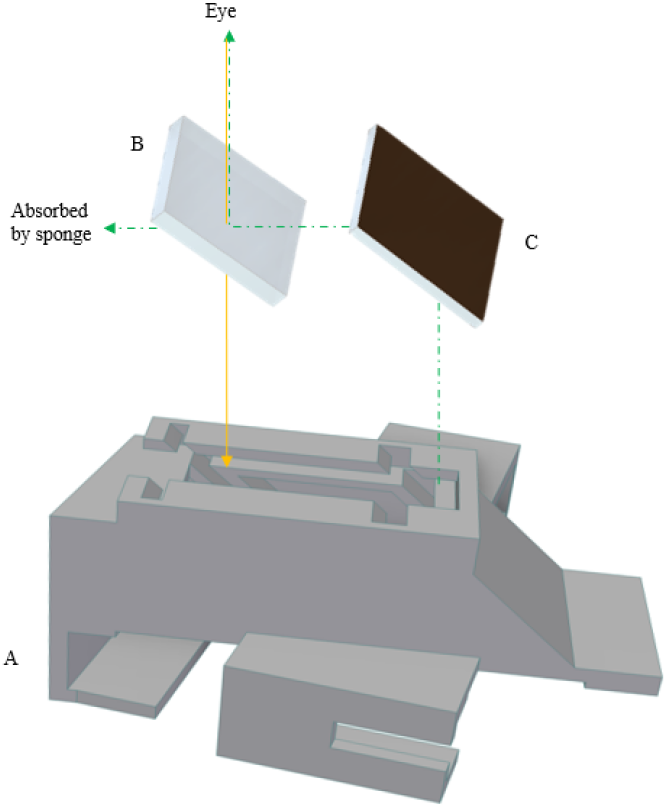
Desgin of GSoP prototype. (A) The 3D model of the case, (B) Beam splitter, (C) Refrective mirror

The prototype depicted in Fig. 2 is specifically designed for the Redmi Note-11 smartphone. However, the design principles are adaptable and can be modified to accommodate other smartphone models, ensuring broader compatibility and accessibility.

**Fig. 2.**
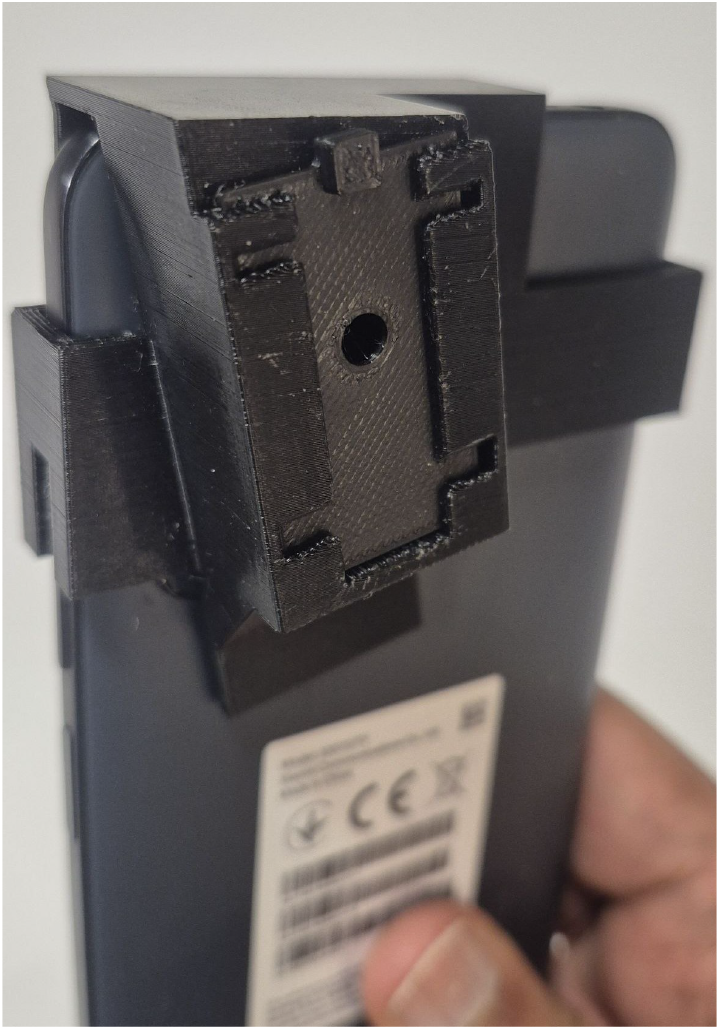
The GSoP adapter attached with the smartphone.

The key optical elements on the designed GSoP device include:

- Transparent Paper: Reduces the intensity of the smartphone flashlight to avoid overexposure. It is placed on top of the flashlight.
- Reflective Mirror: Directs the flashlight beam toward the semi-transparent beam splitter.
- Beam Splitter: Diverts light toward the patient’s eye while allowing light reflected from the eye to pass through to the smartphone camera.
- Black Sponge: Absorbs excess reflected light that passes through the beam splitter, reducing glare.

When properly assembled, the GSoP adapter measures approximately 64 × 81 × 31 mm and weighs around 48 grams, making it lightweight and portable. Despite its low production cost of under C10, the device is designed to be durable and contain lesser materials. The ONH region is displayed on the smartphone screen for examination. To facilitate patient positioning, we developed a custom chin and headrest prototype, as shown in Fig. 3. The main structure of the headrest is constructed from aluminum metal frames, while 3D-printed plastic components are used for the sections that come into contact with the patient’s chin and head. This design ensures stability during video recording and ONH assessment, minimizing distortions and providing a clearer, less-distorted view of the target region.

**Fig. 3.**
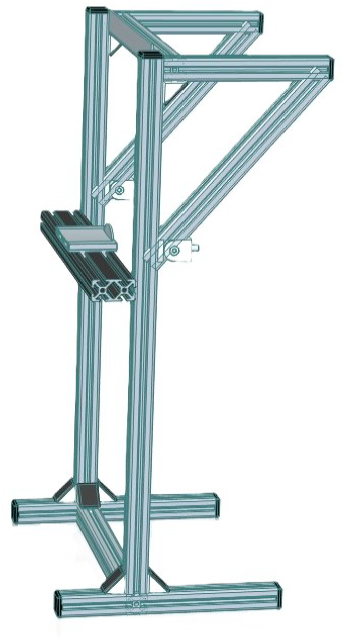
A Chin and Headrest prototype for fundus examination,

### B. Video Recording Protocol

Clinical testing of the GSoP device was conducted at the Ophthalmology Department of Jimma University Medical Center (Jimma, Ethiopia) following approval from the Institutional Review Board (IRB) of the University. The study adhered to the principles of the Declaration of Helsinki and included both healthy participants and those diagnosed with glaucoma. Written informed consent was obtained from all participants prior to the testing. The study involved a comparative analysis of videos captured using the GSoP and oDocsnun ophthalmoscope devices. Both devices were used under controlled conditions to record videos of the participants’ optic disc (OD) regions, enabling a direct comparison of image quality from same subject.

### C. Video Acquisition

An Android application was developed to manage patient information, record videos, and store data locally on the smartphone. The application is compatible with both the GSoP and oDocs-nun ophthalmoscope devices, ensuring seamless video acquisition and data storage. The image capturing process is based on the principles of direct ophthalmoscopy, with the examiner positioning the device at an optimal distance to capture the target region clearly, using the chin-and-headrest shown in Fig. 3.

The video recordings of 10-15 seconds per eye were captured using a Redmi Note-11 smartphone equipped with a 50MP camera, recording at 30 frames per second. The video acquisition process involved the following steps:

1. Obtain informed consent from the participant.
2. Position the patient using the chin and headrest.
3. Launch the Android application and prepare the device.
4. Record videos of each eye by placing the smartphone around 1 cm away form the eye.
5. Save the recordings to the smartphone’s local storage for further analysis. The recorded videos are then transferred securely to a location where only the research team of this study has access.

## IV. Results and Discussion

The primary objective of this study is to develop an affordable fundus imaging device capable of supporting glaucoma screening. The region of interest (ROI) for this research was the ONH, a critical area for detecting early signs of glaucoma. Videos of the ONH were recorded using the GSoP and oDocs-nun ophthalmic devices from dilated pupils, and sample frames were analyzed in terms of their FOV and image quality.

### A. Image Quality and Reflections

As shown in Fig. 4, images captured with the GSoP device exhibited circular glare artifacts, primarily caused by the reflection of the smartphone’s flashlight on the eye’s surface. Additionally, sharpness-related noise reduced the clarity of the ONH region, posing challenges for diagnostic interpretation. Although integrating a black sponge into the 3D-printed case helped reduce excess reflection, it did not fully eliminate glare. These limitations reflect the balance between affordability and image quality, as the GSoP was designed to be a low-cost, accessible alternative rather than a direct replacement for high-end ophthalmic devices.

**Fig. 4.**
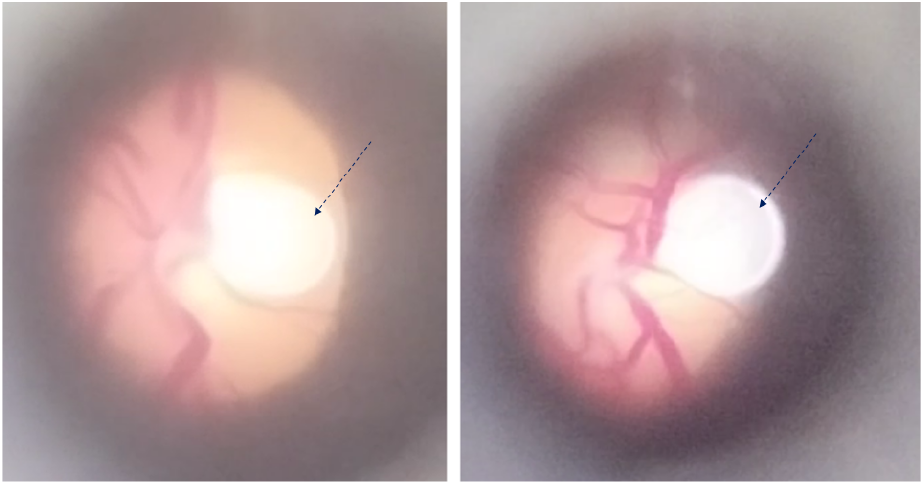
Sample images manually selected from videos recorded using GSoP adapter. It shows the glare (with the black arrow) from the smartphone’s flashlight. The visible retinal area is the boundary of the OD

In contrast, videos recorded with the oDocs-nun ophthalmoscope, as shown in Fig. 5, displayed significantly fewer artifacts, including reduced glare and noise. Its advanced optical design and integrated noise reduction mechanisms minimized glare and noise, providing higher image clarity. However, these enhancements increase its production cost, making the device less accessible for widespread use in resource-limited settings.

**Fig. 5.**
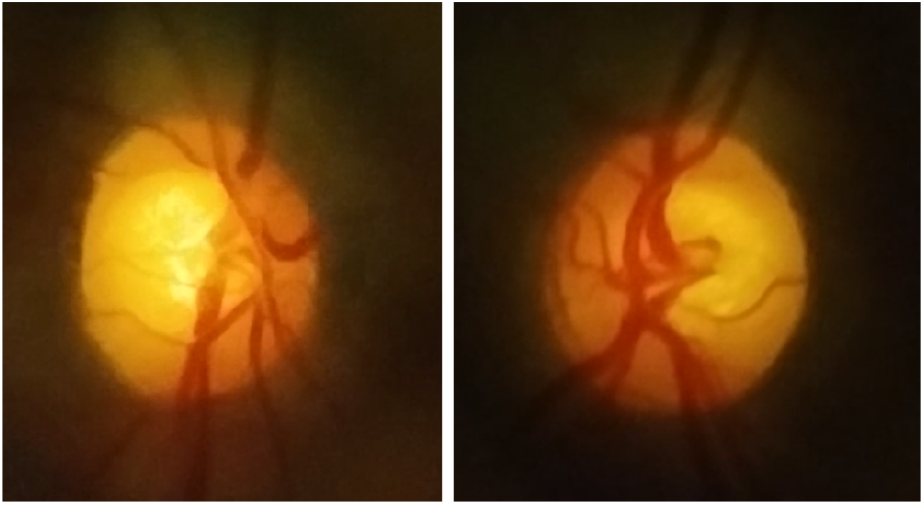
Sample images manually selected from videos recorded using oDocs-nun ophthalmoscope

While the oDocs-nun offers higher image quality, the GSoP remains a viable option for screening purposes, as it can still capture diagnostically relevant ONH features. When combined with video-based analysis and post-processing techniques, the GSoP can help compensate for its optical limitations, ensuring that essential retinal assessments remain feasible. The comparison highlights the trade-off between cost-effectiveness and image quality, with the GSoP prioritizing affordability while maintaining sufficient diagnostic utility.

Future improvements for the GSoP device could include the implementation of post-processing techniques, such as denoising algorithms and glare reduction models, to enhance image quality. Additionally, incorporating cost-effective anti-reflective coatings or optimizing the optical alignment could further mitigate glare without significantly increasing production costs. These enhancements would improve the usability of the GSoP, ensuring it remains a practical and affordable tool for glaucoma screening in primary healthcare settings.

### B. Field of View

The GSoP device had a smaller FOV of up to 20 degrees compared to the oDocs-nun ophthalmoscope. Despite this limitation, the GSoP effectively captured the complete boundary of the OD in a single frame, as shown in Fig.4. This is particularly relevant for glaucoma screening, as early indicators such as changes in the cup-to-disc ratio are often observed in the OD. The ability of the GSoP to capture the ONH parts on multiple video frames ensures the visibility of critical glaucoma indicators.

Although the smaller FOV restricts the GSoP’s ability to capture peripheral retinal details, it does not hinder its primary purpose of ONH examination. Expanding the FOV in future versions of the device, while maintaining affordability, could enhance its utility for broader diagnostic applications, including the screening of other retinal diseases.

### C. Video Recording and Frame Analysis

The GSoP relies on video recording over a duration of 10–15 seconds, enabling the capture of multiple frames and increasing the chance of obtaining diagnostically relevant images. This video-based approach not only compensates for challenges such as reduced FOV and occasional image artifacts but also allows the examiner to select the clearest and most informative frames for analysis. For example, Fig. 6a-c demonstrates how sequence of frames collectively provide comprehensive information about the ONH, which would be difficult to capture in a single image. This method ensures that diagnostically valuable data can still be extracted, even under suboptimal imaging conditions.

**Fig. 6.**
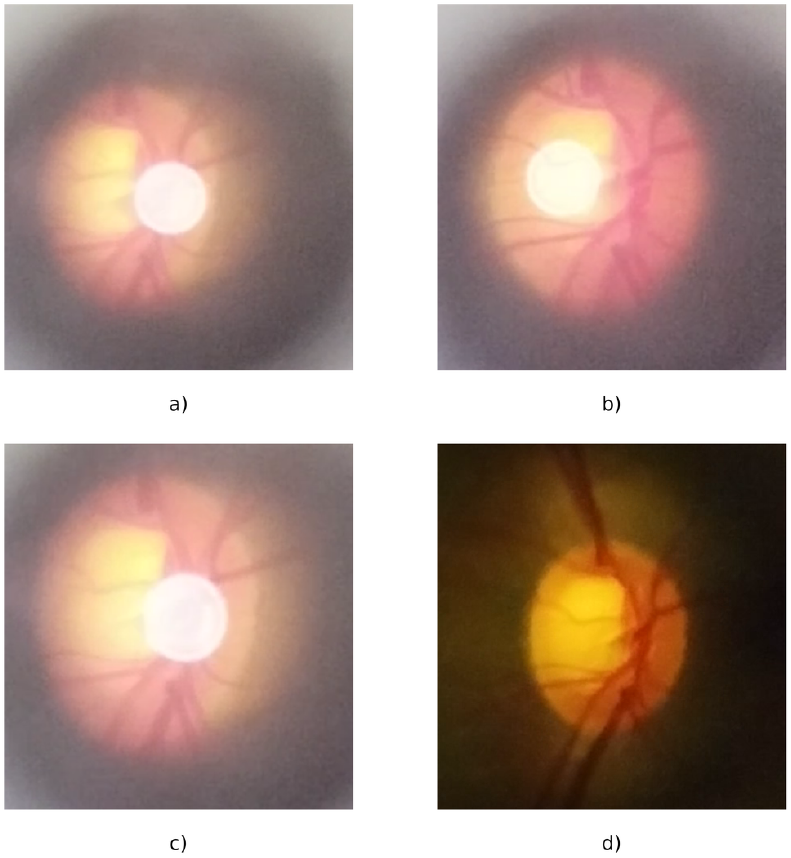
Glaucomatous sample images from GSoP (a-c), showing areas obscured by the reflection in one frame are visible in others. The image from oDocs-nun ophthalmoscope (d) is also captured from the same eye

The visibility and features of the ONH, including its shape, color, and boundary definition, are critical for assessing glaucoma indicators. Fig.6 and Fig.7 display sample images of glaucomatous and healthy eyes, respectively, captured with both the GSoP and oDocs-nun devices. While the oDocs-nun ophthalmoscope produced higher-quality images (Fig.6d and Fig.7d), the GSoP’s video-based approach allowed for capturing multiple perspectives of the ONH. Despite noticeable artifacts, such as circular glare in frames from the GSoP recordings (Fig.6a–c and Fig.7a–c), the variation in pixel positions across frames ensures that regions obscured in one frame may be visible in others. With inclusion of automated algorithms, the ONH features across multiple frames can be analyzed.

**Fig. 7.**
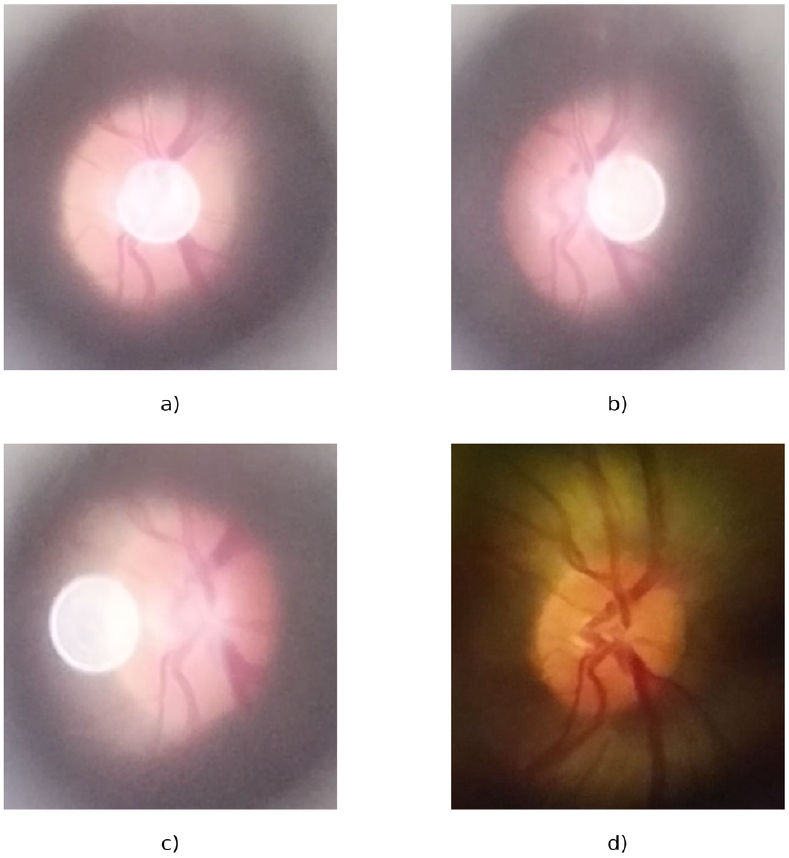
Healthy ONH images captured with GSoP (a–c) and oDocs-nun (d). GSoP’s multi-frame capture yields comprehensive details comparable to a single high-quality oDocs-nun image

The trade-off between image quality and cost-effectiveness underscores the practicality of the GSoP. By leveraging video recordings, the device compensates for its optical limitations and provides relevant diagnostic information for glaucoma screening. The ability to extract useful insights from multiple frames highlights the GSoP’s potential as an accessible and affordable tool for primary healthcare settings.

## V. CONCLUSIONS

This study demonstrated the design, development, and evaluation of the GSoP, an affordable smartphone-based fundus imaging device specifically designed for glaucoma screening and educational applications. The findings highlight the device’s capability to capture diagnostically relevant ONH images, despite challenges such as glare and noise. By employing a low-cost design, the GSoP addresses critical barriers posed by traditional fundus cameras, including high costs, and limited accessibility in resource-constrained settings. The device’s portability, affordability, and ease of use make it an invaluable tool for primary healthcare facilities and outreach programs.

One of the GSoP’s most notable advantages is its affordability, with a production cost of less than C10. This positions the device as a highly economical alternative to existing smartphone-based ophthalmoscope devices, which, though marketed as affordable, often remain out of reach for resource-limited healthcare systems. By significantly reducing production costs and simplifying the design, the GSoP effectively addresses the financial and operational challenges that hinder the widespread adoption of fundus cameras in low-resource environments. This affordability enables the GSoP to be deployed more broadly for glaucoma screening campaigns, educational outreach, and primary care in underserved communities.

Future work will focus on improving image quality by incorporating software-based enhancements, such as denoising algorithms and glare reduction techniques. Additionally, integrating artificial intelligence for automated glaucoma screening will further enhance the device’s diagnostic utility, reducing reliance on specialist interpretation and making it even more practical for primary healthcare settings. These advancements have the potential to significantly expand the GSoP’s impact on global eye health and strengthen its role as a cost-effective solution for combating preventable blindness.

## Data Availability

All data produced in the present study are available upon reasonable request to the authors

